# Clinical trials in depression: Integrated collection across EU and US registries

**DOI:** 10.1101/2025.10.10.25337719

**Authors:** Kate Stewart, Louise Schindler, Robert Bevan, Samrina Rehman, Matthew H. Iveson, Andrew M. McIntosh, Naomi R. Wray, AMBER Research Team, Cathryn M. Lewis

## Abstract

Depression affects millions worldwide with both pharmacological and psychological therapies widely applied, both with limited treatment success. Many clinical trials have been undertaken to test and compare treatments for depression. In collaboration with Medicines Discovery Catapult (MDC), we developed a comprehensive database of depression-related clinical trials, including information on the availability of additional biological samples such as blood or DNA. We systematically extracted structured and unstructured data from two major clinical trial registries: the US (https://clinicaltrials.gov/, using the Aggregated Analysis of ClinicalTrials.gov (AACT) database) and the EU Clinical trials register (https://www.clinicaltrialsregister.eu/). Summary data from clinical trial records and resulting publications were extracted on interventions, conditions, drugs, sample sizes, trial phase, status, and start and end dates, participant demographics, and sponsors. To identify trials likely to include blood samples or genetic information, we applied a semantic similarity approach using vector-based natural language processing to score trial records based on textual indicators. This methodology prioritised trials most likely to contain genetic information by measuring similarity between predefined query terms and clinical trial text fields.

We identified 8,853 unique clinical trials registered between 1987 and 2024, with the majority (86%) from the AACT database. In total, 3,659 (41%) trials involved a drug intervention, 1,160 of which were testing antidepressants. Selective serotonin reuptake inhibitors (SSRIs) were the most commonly tested drug class (n=899), with escitalopram the most frequently studied drug (n=322). Other trial interventions included behavioural interventions and digital devices. SSRI trials peaked around 2009, while most trials since 2015 have been of NMDA receptor antagonists such as ketamine. Trial sponsors included universities, healthcare organisations, and pharmaceutical companies, with the latter sponsoring a higher proportion of drug intervention trials. Vector scores prioritised trials likely to have biological samples or genetic data, and initial validation confirmed the accuracy of these indicators. Participant demographic data and final sample size were incomplete for many trials.

This comprehensive resource of clinical trials in depression provides a valuable overview of the landscape of trials performed since 1987. The dataset will be shared as an open-access repository on publication. It will enable researchers to identify clinical trials with genetic samples to expand pharmacogenetic studies, ultimately working toward the goal of personalised antidepressant prescribing.

**Plain language summary:** Depression affects millions worldwide. Both medications and psychological therapies are used to treat depression, but the rates of those who respond to treatment are low. Many clinical trials have tested and compared treatments for depression. Pharmacogenetics, the study of how a person’s genetics can influence how they respond to a particular drug, is one approach which may improve treatment by using genetics to guide treatments for individual patients.

In collaboration with Medicines Discovery Catapult (MDC), we developed a comprehensive collection of depression-related clinical trials. We extracted data from two major clinical trial registries in the US and the EU. We summarised information about the interventions tested; conditions and diagnoses studied alongside depression; phase, status and start and end dates of the trials; information about trial sponsors; and demographics information about trial participants. We also applied informatics methods to identify which trials were likely to include genetic information or blood samples.

We identified 8,853 unique clinical trials registered between 1987 and 2024, with most (86%) from the US register. Over 40% of clinical trials included a drug intervention, with around 13% testing antidepressants. Other trials included behavioural interventions and digital devices. Trial sponsors included universities, healthcare organisations, and pharmaceutical companies. Information about participant demographics and final sample sizes were incomplete for many trials.

This comprehensive resource of clinical trials in depression provides a valuable overview of the landscape of trials performed since 1987. It will allow researchers to identify relevant clinical trials to expand pharmacogenetic studies and work toward the goal of improving treatments for depression.

## Introduction

Depression poses a significant global challenge, affecting millions of individuals worldwide and contributing substantially to the global burden of disease. It has become an increasing problem, rising to the second highest contributor to years lived with disability globally.^1^ In the UK, annual antidepressant prescriptions increased from 36 million to 71 million between 2008 and 2018.^2^ This trend highlights the critical need for improved strategies to identify and tailor effective depression treatment.

Treatment for depression includes both psychological and pharmacological therapies, both of which have been used over the last three decades. Psychological therapies are widely available, ranging from self-referral to talking therapies in the UK NHS, to community-based approaches such as the Friendship Bench, an evidence-based intervention developed in Zimbabwe to bridge the mental health treatment gap.^3^ Selective serotonin reuptake inhibitors (SSRIs) are widely prescribed, and the recent development of newer N-methyl-D-aspartate (NMDA) receptor antagonists, such as ketamine, has attracted considerable interest. Clinical trials form the evidence base for both pharmacological and psychological depression treatments,^4^ with thousands of trials being performed across treatment modalities.

Despite decades of work to improve options for treating depression, treatment response rates for depression remain low. For example, only approximately one third of patients reach remission during a treatment period with antidepressants^5^ and rates are similar between pharmacological and psychological therapies.^6^ There are currently few predictors of treatment response to direct personalized prescribing, and many patients undergo trial-and-error prescribing to find a suitable treatment.

Pharmacogenetics offers a promising avenue to improve depression treatment, using genetic influences on drug response to personalise depression treatment. Clinical guidelines for using cytochrome P450 metabolising status when prescribing SSRIs recommend dose reductions and alternative drugs where appropriate, but much remains unknown about the impact of genetics on treatment response and adverse events.^2^

Research in pharmacogenetics faces several challenges; studies require detailed, longitudinal data collection to accurately assess treatment response, and large sample sizes across studies are needed to identify modest genetic effects. Genome-wide association studies have shown moderate heritability of antidepressant response, underscoring the potential to identify further pharmacogenetic variants.^7^

Clinical trials represent a valuable resource for addressing these limitations. Trials systematically assess the efficacy and safety of antidepressant interventions over time, generating rich clinical datasets that may include genetic data, or samples from which genetic data can be generated. Leveraging these clinical trials could greatly enhance pharmacogenetic research in depression. While individual clinical trials tend to be relatively small, they can be systematically combined to create more robust statistical estimates. However, this approach first requires a comprehensive catalogue of eligible trials which might be suitable for combination and analysis.

To more fully exploit the value of clinical trials in depression for pharmacogenetic studies, we generated a comprehensive data resource of clinical trials performed in depression. Using clinical trial registries in the US and the EU, we mapped information on trials, building a structured open access dataset, using text mining and natural language processing to enrich data from the online registries. This AMBER-MDC resource compiles detailed information, including patient demographics and trial characteristics, to facilitate a wide range of subsequent analyses. Trials across treatment modalities are included, spanning pharmacological, behavioural, digital and other interventions. This systematic approach enables downstream research to better understand antidepressant action, particularly the identification of genetic determinants of treatment response.

## Data summary

### Development of clinical trials dataset

We accessed information on clinical trials with drug interventions registered on ClinicalTrials.gov and EU Clinical Trials Register, as these represent the largest publicly accessible registries of clinical trials globally. Data from these sources were processed using different approaches due to varying database structures and available metadata and then combined into a single dataset (Figure 1).

**Figure 1.**
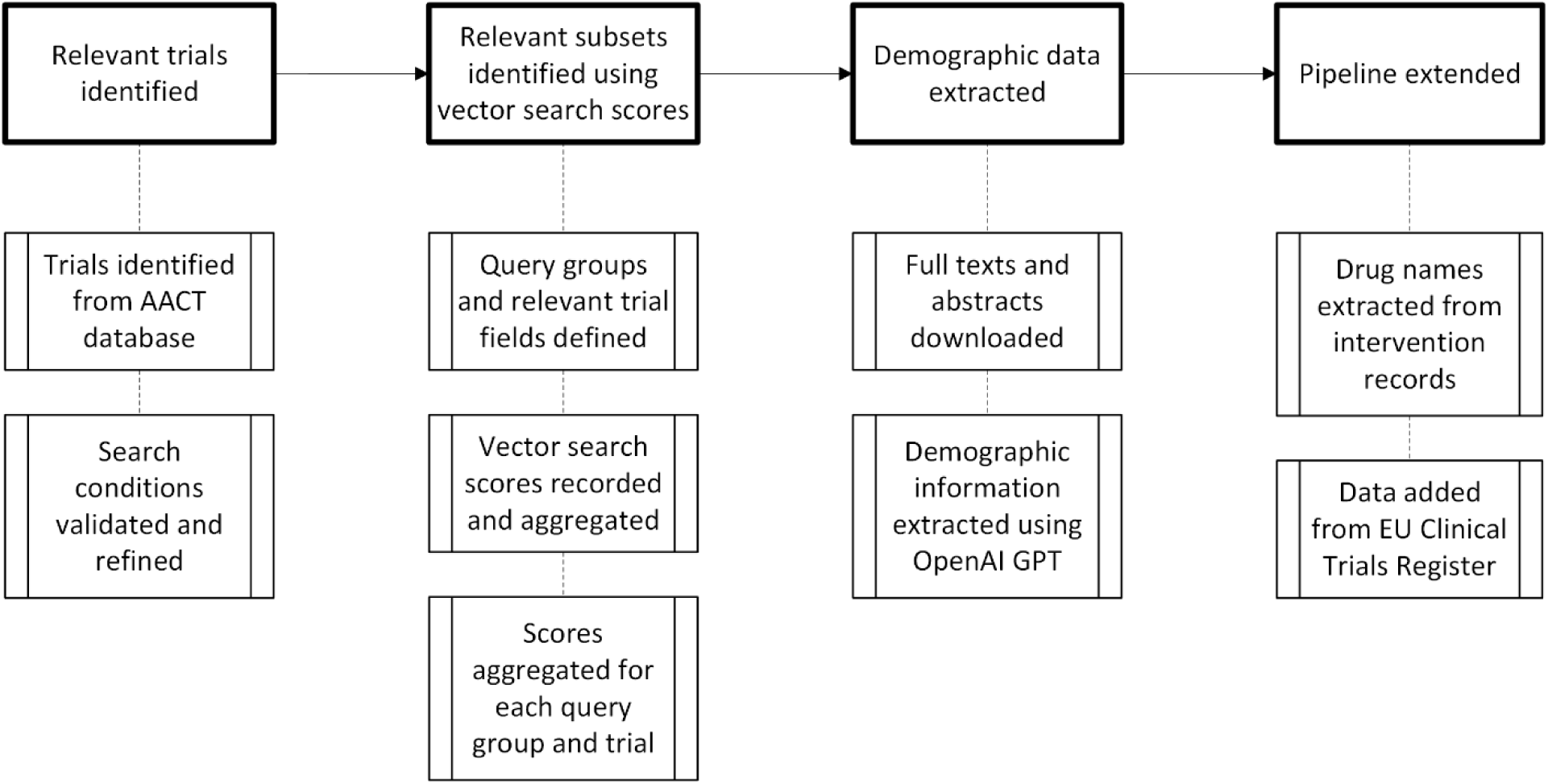
Flow chart showing how clinical trial data, including trial characteristics, demographics and intervention details, were identified and extracted.

### ClinicalTrials.gov data collection

#### Trial identification and filtering

The Aggregate Analysis of ClinicalTrails.gov (AACT) database is a publicly available relational database, updated daily, that lists all trials registered on ClinicalTrials.gov. We included all trials registered which met the criteria from the database launch date in 1987 through June 2024 to build an initial list of depression clinical trials. Depression trials were identified by matching the conditions table to a regular expression based on the following MeSH terms: *depression; depressive disorder; depressive disorder, major; depression, postpartum; depressive disorder, treatment-resistant*. The set of conditions associated with the trials returned by the query were manually validated and irrelevant conditions discarded. This refined condition set was used as a filter to identify depression trials, including those investigating comorbid conditions alongside depression (e.g. cancer and depression). Data extraction was performed in June 2024.

#### Vector-based search for biological samples

To identify depression trials likely to contain biological samples and omics datasets, we developed a vector-based search method. Since the AACT database lacks explicit metadata for biological data collection, we used natural language processing tools to extract this information from free-text fields within clinical records. First, we defined a small set of representative query phrases for each category of interest (e.g., ‘blood sample’, ‘genomics’) and selected the free-text fields in AACT most likely to mention specimen collection (e.g., ‘outcome measures’) (Supplementary Table 1). Query terms and trial field text were converted to a semantic vector with OpenAI’s text embedding API, and cosine similarity was calculated between each query vector and field vector. For fields with multiple entries (e.g., several outcome measures), each entry was scored individually, and the highest score was retained for that field. Scores were then aggregated by taking the maximum similarity score within each query-field combination and further aggregated at the query group level. This yielded a single similarity score per category per trial (e.g., ‘blood sample score’, ‘genomics score’). This approach enabled systematic identification of trials with potential biological sample availability from unstructured text data across the depression trial dataset.

#### Demographic data extraction

To populate demographic variables, we built a multi-step text-mining pipeline that draws on both ClinicalTrials.gov content and linked literature. First, we retrieved each trial’s free-text eligibility criteria, study protocols and publication links directly from AACT. Publication links were used to get PubMed IDs for relevant trials, including only publications assigned as “result” and “derived” in AACT, excluding “background” publications. PubMed Central and PubMed APIs were used to download full text publications. Where full text publications were unavailable (e.g. behind a paywall, not deposited in PubMed Central), abstracts were downloaded from PubMed. Up to five publications per trial were processed, prioritising those with available full text and excluding any predating trial initiation. Two tailored OpenAI GPT prompts were then applied: one parsed publication text, the other eligibility criteria. From publications we extracted total sample size, male and female counts, ethnicity counts, and age and BMI statistics (minimum, maximum, mean, standard deviation). From eligibility criteria we captured the planned sample size plus minimum and maximum age and BMI. Ethnicity labels returned by GPT were manually mapped to the NIH standard set,^8^ with unmapped values re-coded as “Other”. Where multiple sources reported the same field for a trial, we assigned the modal value; if no mode existed, we used the mean. Both aggregated and raw extracted data are provided at https://kcl.figshare.com/articles/dataset/_b_Clinical_trials_in_depression_Integrated_collection_across_EU_and_US_registries_b_/30217441/0. Finally, to standardise pharmacological information, GPT was used to isolate drug names from the free-text intervention field, then each drug was queried against the PubChem API to obtain its PubChem CID, InChI string, and WHO ATC antidepressant classification. No information on trial results was extracted.

### EU Clinical Trials Register data collection

#### Web scraping and data mapping

To expand our dataset coverage, we scraped trial records for depression from the EU Clinical Trials Register using the MeSH terms as above. Unlike ClinicalTrials.gov, no publicly available relational database exists for the EU Clinical Trials Register. Fields were mapped to the corresponding fields in ClinicalTrials.gov where possible. The records obtained from the EU Clinical Trials Register are sparser than those from the AACT database due to different database schemas. EU Clinical Trials Register records do not include links to related publications, so demographic information extraction relied solely on data present within the trial records.

### Dataset integration and standardisation

The extracted clinical trial data exhibit substantial heterogeneity due to incomplete records, use of free text within standardised trial register fields, and structural differences between clinical trial registries. To address these limitations, we implemented standardisation procedures, documented at https://kcl.figshare.com/articles/dataset/_b_Clinical_trials_in_depression_Integrated_collection_across_EU_and_US_registries_b_/30217441/0 and on GitHub at https://github.com/louisesophieschindler/Clinical-trials-in-depression and https://github.com/katemstewart/Clinical_Trials_in_Depression. Both raw and processed datasets are provided in the repository.

#### Derived dataset creation

Additional processed datasets were derived for specific analyses, focussing on trials of pharmacological interventions, extracting trials for antidepressants and other drug interventions. Data processing focussed on two domains from the complete dataset (All Clinical Trials [ACT]): detailed trial records and detailed intervention records. Detailed trial records were filtered by drug intervention status to identify trials including drug interventions (Drug Intervention Trials [DIT]). From detailed intervention records, we extracted trials with established antidepressant classes (Antidepressant Trials [ADT]), excluding other drugs (e.g., immunotherapy, chemotherapy, vitamins) and chemical compounds in development, which are listed only as pharmaceutical compound number prefixes (e.g., GSK561679, JNJ-54175446). We separately extracted trials involving NMDA receptor antagonists, cannabinoids, or psychedelics (Emerging Depression Intervention Trials [EDIT].). For both extractions, drug classes were standardised and active ingredients classified by generic drug name. Finally, we created a comprehensive dataset combining all depression-related drugs (All Depression-Focused Drug Trials [ADDT]): antidepressants, NMDA receptor antagonists, cannabinoids, and psychedelics (Figure 2).

**Figure 2.**
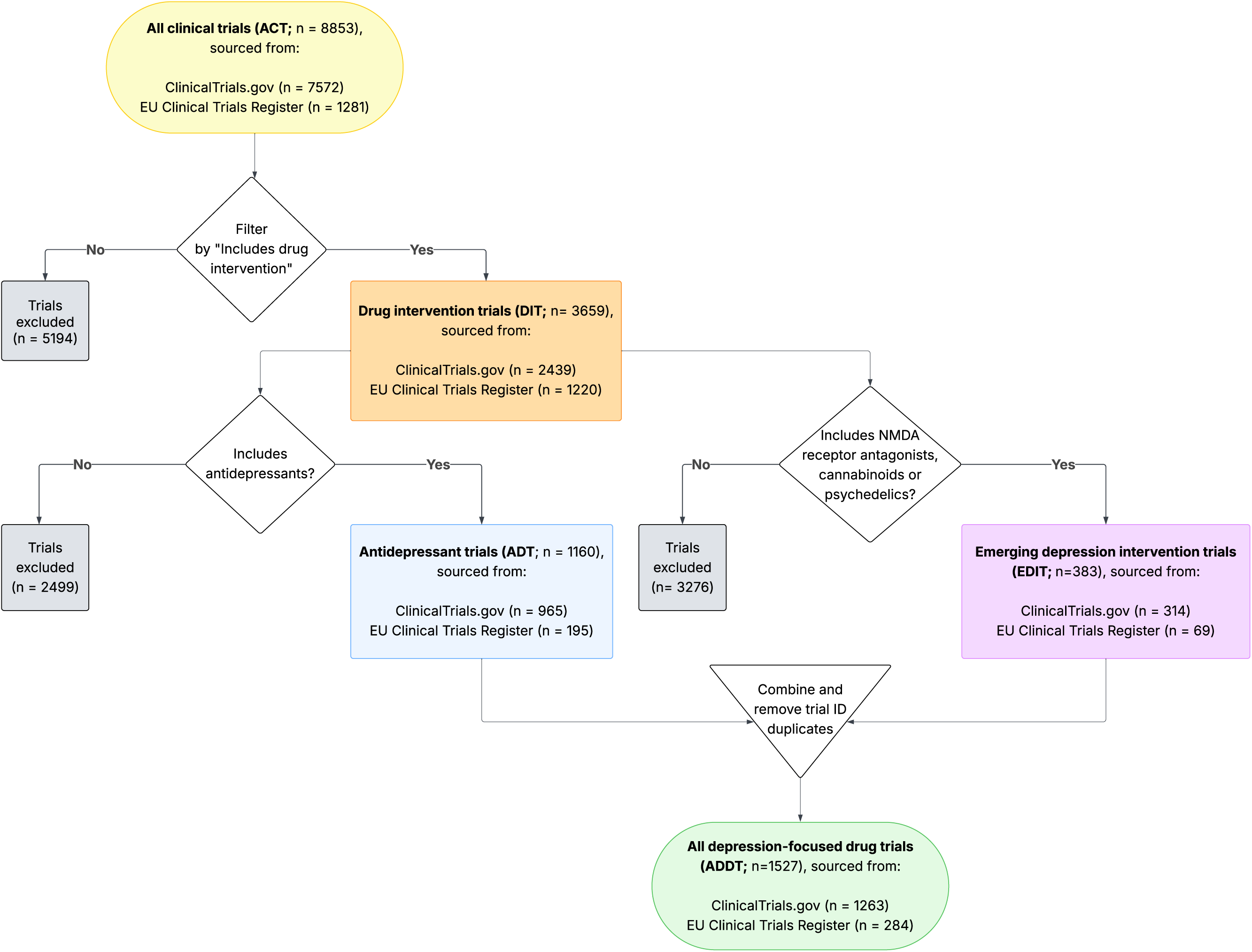
Flow chart showing how depression clinical trials across ClinicalTrials.gov and EU Clinical Trials Register were processed to extract trials, and trial arms, with antidepressant and other drug treatment for depression.

#### Sponsor classification

Sponsors of clinical trials were categorised into three groups, and summarised:

- Healthcare industry: pharmaceutical company, device company or contract research organisation (CRO);
- Academic and public sector: university, government organisation, collaborative group, hospital, non-profit organisation or individual; and
- Digital health sector: companies developing and delivering digital solutions such as mobile applications, AI technologies and other digital therapeutics and interventions.

#### Condition variable cleaning

The dataset comprised 8,853 individual trial records containing a free-text description of study conditions. Each free-text entry was extracted, converted to lowercase, trimmed of leading and trailing whitespace, and cleaned of punctuation, bracketed expressions, and connector words using compiled regular expressions. Entries exceeding 200 characters or containing study-related keywords (e.g., “aim”, “objective”) were flagged as “long descriptions” (n = 128) and excluded from further tokenisation. The remaining 8,712 trial records were split on semicolons into discrete terms.

These terms were manually reviewed and categorised. Only terms appearing more than once were classified. Two lookup tables were created for categorisation. The first contained 14 top-level categories and 39 subcategories covering 507 unique clinical terms (Table 1). For example, unique clinical terms “anhedonia”, “currently depressed”, and “depressive disorder” were grouped under the subcategory “depression” within the top category “mental & behavioural disorders”. The second lookup table linked 89 unique methodological terms to metadata labels describing study design. Where appropriate, terms were assigned to more than one category (e.g., “postpartum depression” was assigned to “depression” and “female & perinatal”).

**Table 1.**
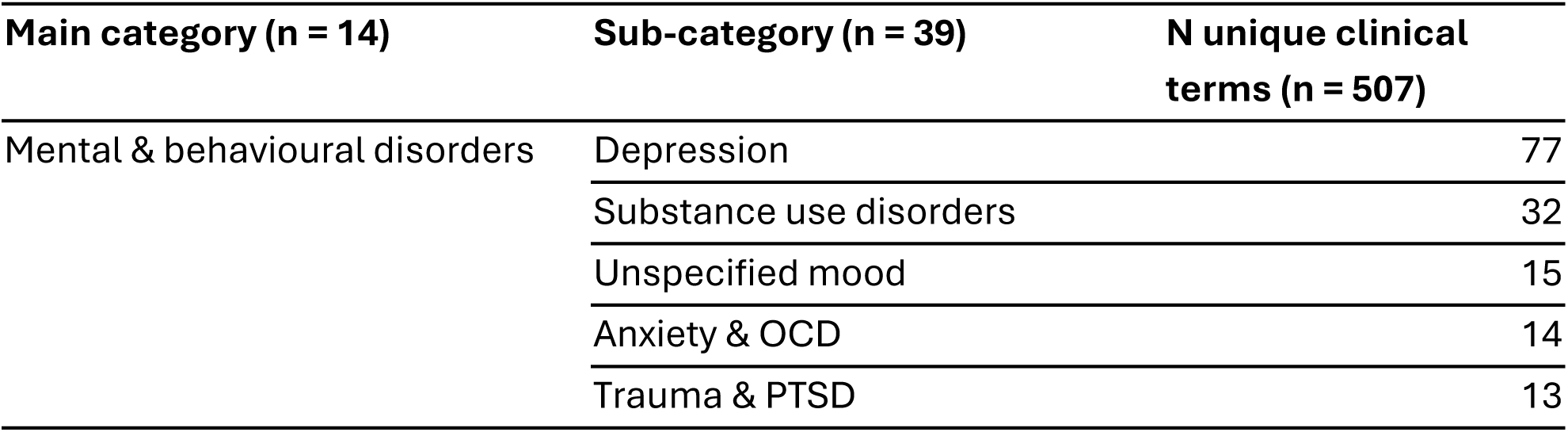
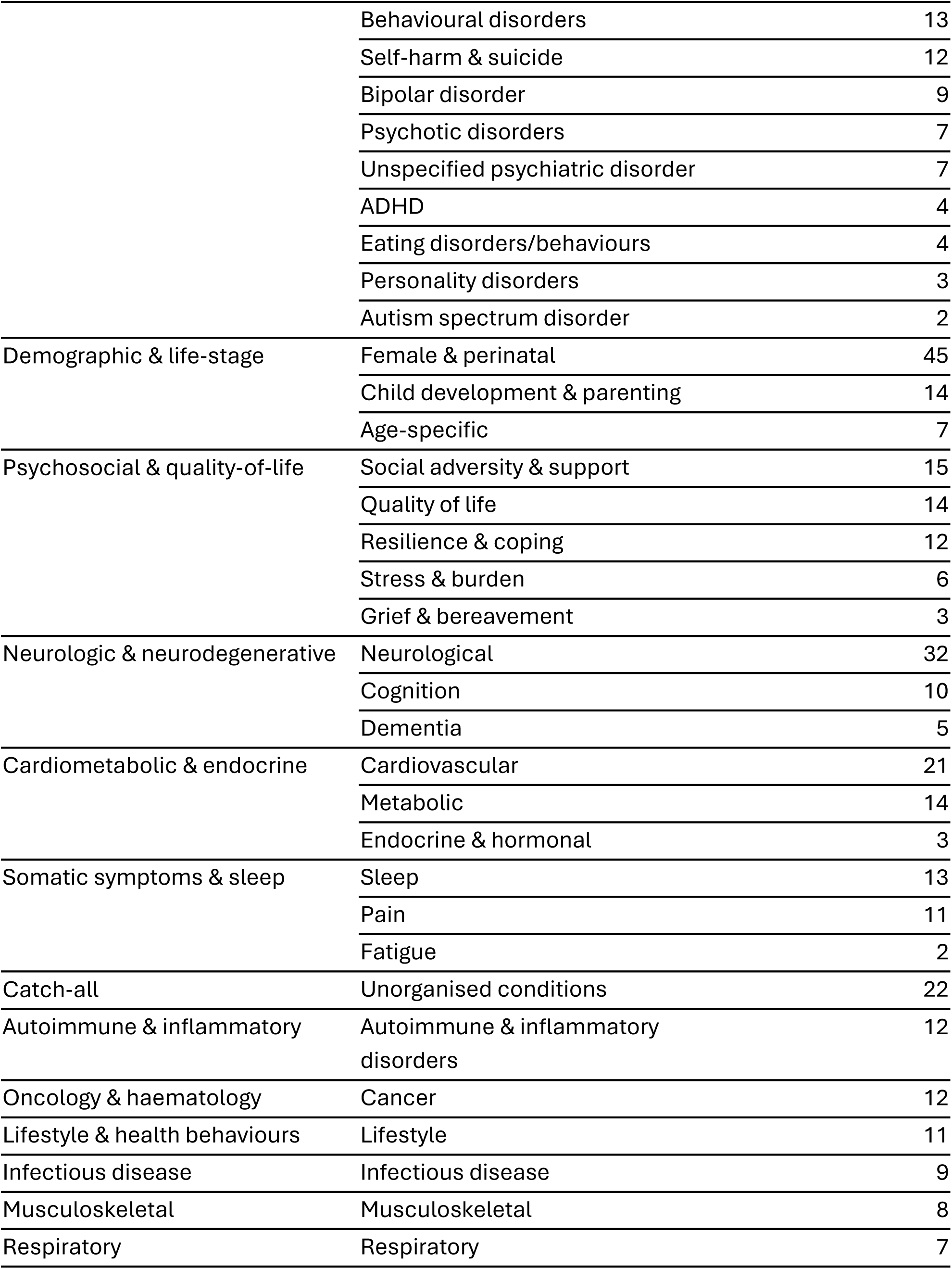
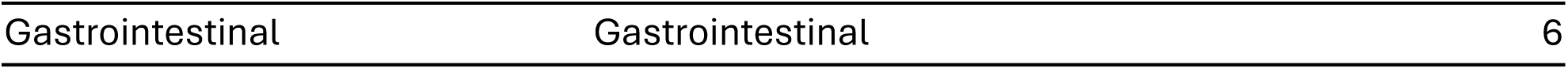
Top categories and sub-categories, including the number of unique clinical terms per sub-category.

Rows containing any terms for a clinical condition were assigned to the “conditions” category. Rows containing only metadata terms were assigned to “metadata”, and rows with no matching terms were labelled “none”. Of the 8,712 trial records processed, 8,449 (97.0%) were assigned to one or more clinical condition labels, 118 records (1.2%) contained only metadata labels, and 145 records (1.7%) had no matches. The fully annotated dataset, enriched with category assignments and unmatched phrase lists, was then exported for downstream analysis.

### Final Dataset Structure and Composition

The final dataset comprises these nine primary data tables: trial summary, trial details, PICO framework (Patient/Population, Intervention, Comparator, Outcome), intervention summary, intervention details, search terms, demographic summary, demographic details, and publications. We have also added domains for antidepressant interventions (ADT), other pharmacological interventions (EDIT), and all pharmacological interventions (ADDT; Figure 2). Further details on the structure of each table, including variable counts and trial coverage, is given in Table 2.

**Table 2.** Summary of datasets, with details of variable and inclusion criteria. Shading aligns with Figure 2.

| Data Table | Variables | Records | Trials | Description |
| --- | --- | --- | --- | --- |
| Trial Summary | 35 | 8,853 | 8,853 | **Core trial metadata:** Identifiers, source registry, drug intervention indicators, and enrolment data, demographic summaries (including trial sample sizes), biological sample availability scores. |
| Trial Details | 53 | 8,853 | 8,853 | **Comprehensive trial information.** extending summary data with interventions, conditions, MeSH terms, study phase, status, dates, and sponsorship. Includes standardised ethnicity classifications mapped to NIH categories. |
| PICO | 21 | 81,551 | 8,853 | **Trial data in PICO format:** Population, Intervention, Comparison, Outcomes. Includes all trial outcomes with primary and secondary designations. |
| Intervention Summary | 3 | 15,601 | 8,853 | **Interventions:** categorised by type (drug, behavioural, device, etc.). |
| Intervention Details | 7 | 15,926 | 8,853 | **Detailed drug interventions:** trials with multiple drug interventions are separated into individual drug records. Includes PubChem identifiers, InChI strings, and WHO ATC classifications. |
| Vector Search Results | 54 | 8,853 | 8,853 | **Vector search scores:** all queries, in all query groups, for each trial. |
| Demographic Summary | 70 | 8,853 | 8,853 | **Demographic statistics** per trial including modal, mean, minimum, and maximum values for trial sample size, age, BMI, gender, and ethnicity. |
| Demographic Details | 22 | 11,629 | 8,853 | **Demographic extractions** from each source for each trial, including eligibility criteria and linked publications. |
| Publications | 4 | 18,228 | 3,094 | **PubMed publications:** including indicators of whether demographics were extracted successfully. |
| Drug Intervention Trials (DIT) | 53 | 3,659 | 3,659 | **Drug intervention trials:** subset of trials with pharmacological interventions, containing full metadata. |
| Antidepressant Interventions (ADT) | 9 | 1,722 | 1,160 | **Antidepressant intervention trials:** detailed drug information including intervention type, drug name, PubChem ID, InChI, and ATC classifications. Complex interventions are decomposed into single-drug records. |
| Emerging Depression Interventions (EDIT) | 9 | 656 | 383 | **Trials involving NMDA receptor antagonist, psychedelic, or cannabinoid compounds:** detailed drug information including intervention type, drug name, PubChem ID, InChI, and ATC classifications. Complex interventions are decomposed into single-drug records. |
| All Depression-Focused Drug Interventions (ADDT) | 9 | 2,378 | 1,527 | **Integrated pharmacological dataset:** all drug-based depression treatments, with antidepressants and other drug interventions. |

### Data Analysis

#### Descriptive Analysis

Key trial characteristics were summarised, including intervention type, drug intervention status, trial phase, conditions, and number of drugs per trial. Drug classes and active ingredients for depression treatments were characterised. Drug classes were stratified according to trial start year. Trial demographics were summarised by sample size, age, sex, and ethnicity. Missingness was analysed. Categorical variables were described using frequency distributions and percentages.

#### Vector Search Score Analysis

Vector search score distributions for each query group were visualised using violin plots to assess relative matching likelihood. Precision at k is defined as the number of relevant results retrieved over a fixed number (k) of results retrieved.^9^ Precision at 20 was calculated for the highest and lowest ranking results for a subset of query groups. For five query groups (Biospecimens, Genetics, Proteomics, SSRIs and Antidepressant Response), we extracted the 20 trials with the highest and lowest vector scores. We manually assessed whether each trial gave a positive result for presence of the vector score term (e.g. biospecimens had been collected) and then calculated the proportion of relevant results among the highest and lowest retrieved trials.^5^

#### Software

All natural language processing tasks described were performed using [OpenAI API](https://openai.com/api/) configuration with GPT4 model gpt-4o-2024-05-13, and text embedding model text-embedding-3-large. Data cleaning and analysis were performed using RStudio 2024.12.1+563 "Kousa Dogwood" Release and Python Version 3.9.20. Figure 1 was produced using Microsoft Visio. Figure 2 was produced using LucidChart.

## Data summary

### Overall dataset coverage

We identified 8,853 unique clinical trials registered on either ClinicalTrials.gov or the EU Clinical Trials Register since 1987 to the extraction date of June 2024. ClinicalTrials.gov contributed 7,572 trials (85.5%) and EU trials contributed 1,281 trials (14.5%).

Across all trials (n = 8,853), a similar proportion of trials were at phases II, III, and IV (12-13% each), but trial phase was reported as ‘Not applicable’ for 55.5% of trials. Among drug-intervention trials specifically, Phase II (25.9%) and Phase III (25.1%) were most common. For trials including drugs to treat depression, Phase IV was the single largest category (34.4%).

Of the total number of clinical trials, 5026 (56.8%) were completed and 3094 (34.9%) trials had associated PubMed ids for paper on results (not background trial information) (Table 3).

**Table 3.** Status of 8,853 trials from ClinicalTrials.gov and the EU Clinical Trials Register.

| Status | ClinicalTrials.gov<br>(n = 7572) | % | EU Clinical Trials Register (n = 1281) | % |
| --- | --- | --- | --- | --- |
| Active, not recruiting | 252 | 2.8 | N/A | N/A |
| Completed | 4,365 | 49.3 | 661 | 7.5 |
| Enrolling by invitation | 67 | 0.8 | N/A | N/A |
| Not yet recruiting | 381 | 4.3 | N/A | N/A |
| Recruiting | 1,097 | 12.4 | N/A | N/A |
| Suspended | 29 | 0.3 | N/A | N/A |
| Terminated | 405 | 4.6 | N/A | N/A |
| Unknown status | 768 | 8.7 | 620 | 7.0 |
| Withdrawn | 208 | 2.3 | N/A | N/A |

Only limited information on sample size, or participant demographic information could therefore be extracted, and these data were only available from ClinicalTrials.gov (Supplementary Table 2, Supplementary Figure 1). For example, information on trial sample size was available for only 2218 trials, with the number of male and female participants available for 944 and 1,145 trials, respectively. Data on ethnicity was even more limited, extracted from only 713 trials. Of these, incomplete information was available from each study on ethnicity groups, potentially because of mismatches between given information and the NIH categories. Age ranges for recruitment were available for 5,394 trials for minimum age (mean: 21.48, SD: 12.07), and 3,450 trials for maximum age (mean: 55.7, SD: 20.69), with only 941 trials having extracted data on the mean age of recruited participants (mean: 42.11, SD: 15.63).

### Trial Interventions

Intervention type was pre-defined according to ClinicalTrials.gov protocol definitions. For trials registered on the EU Clinical Trials Register, intervention type was categorised as “Drug” or “N/A”. Of the 8,853 trials in ACT, drugs were the most frequently reported intervention (41.3% of trials; n = 3659). There were 6,751 individual drug instances because multi-arm trials could list more than one active agent, within or across arms. Behavioural interventions were also common (5,019 instances in 3291 trials, 37.2%) as were device-enabled/mobile interventions (1,452 instances in 986 trials, 11.1 %) (Figure 3; Supplementary Table 3).

**Figure 3.**
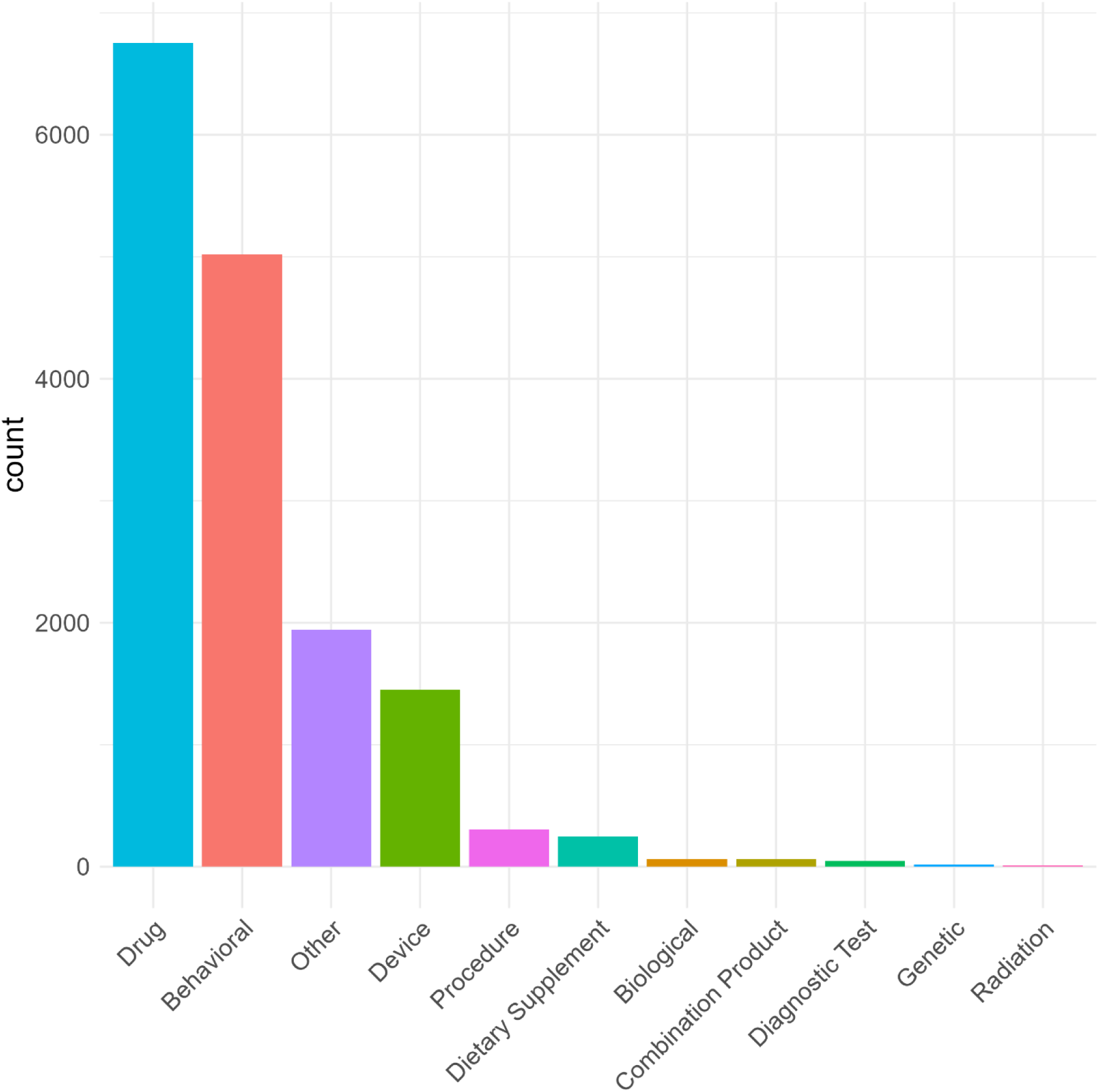
Distribution of trial intervention categories (n=8,853). Many trials had multiple interventions within or across trial arms, (for example a drug intervention with and without psychological treatment).

### Sponsor profile

Most trials were sponsored by academic and other public sector organisations: 78.9% of all trials (6,733), compared with 19.1% (1,630 trials) by healthcare industry and 0.8% (70 trials) by digital health sector companies; the sponsor of 1.2% trials was uncategorised or missing. Among drug intervention trials (DIT; n = 3,659), academic/public sponsors were still the largest group (56.3%, 2,061 trials) but industry participation rose to 38.5% (1,410 trials). In trials investigating drugs for depression (ADDT; n = 1,527), the proportion of academic/public sponsors was 64.2% (981 trials) vs 29.8% (455 trials) for industry (Table 4).

**Table 4.** Distribution of sponsors by trial, and by drug intervention. Drugs to treat depression were specified as antidepressants, NMDA receptor antagonist, psychedelic, or cannabinoid compounds.

| Sponsor type | All trials<br>(ACT; n=8,853) | Drug intervention<br>(DIT; n=3,659) | Drugs to treat depression<br>(ADDT; n=1,527) |
| --- | --- | --- | --- |
| Academic & public sector | 6,733 (76.1%) | 2,061 (56.3%) | 981 (64.2%) |
| Healthcare industry | 1,630 (18.4%) | 1,410 (38.5%) | 455 (29.8%) |
| NA | 415 (4.7%) | 186 (5.1%) | 91 (6.0%) |
| Digital health sector | 70 (0.8%) | 1 (<0.1%) | 0 |
| Unknown | 5 (0.1%) | 1(<0.1%) | 0 |

The largest individual sponsors included large U.S. research hospitals and universities (e.g., Harvard University, Massachusetts General Hospital, Veterans Health Administration) as well as several multinational pharmaceutical companies, the latter mostly for drug-intervention studies (Supplementary Table 4).

### Drug intervention details

Among the 1,527 trials evaluating pharmacological treatments for depression (ADDT), 85.3% included a single drug and 11.1% included two drugs, with the remainder including three or more drugs. Counting each agent separately, the SSRI class was most common (n = 899), followed by SNRIs (n = 367). The most common antidepressant tested was escitalopram (n = 322), followed by sertraline (n = 173) and duloxetine (n = 150) (Figure 4). NMDA-receptor antagonists were also widely tested in clinical trials (n = 536), with ketamine (n = 445) being the most common agent. Psychedelics were included in trials only 78 times, and cannabinoids only 38 times (Supplementary Figure 2).

**Figure 4.**
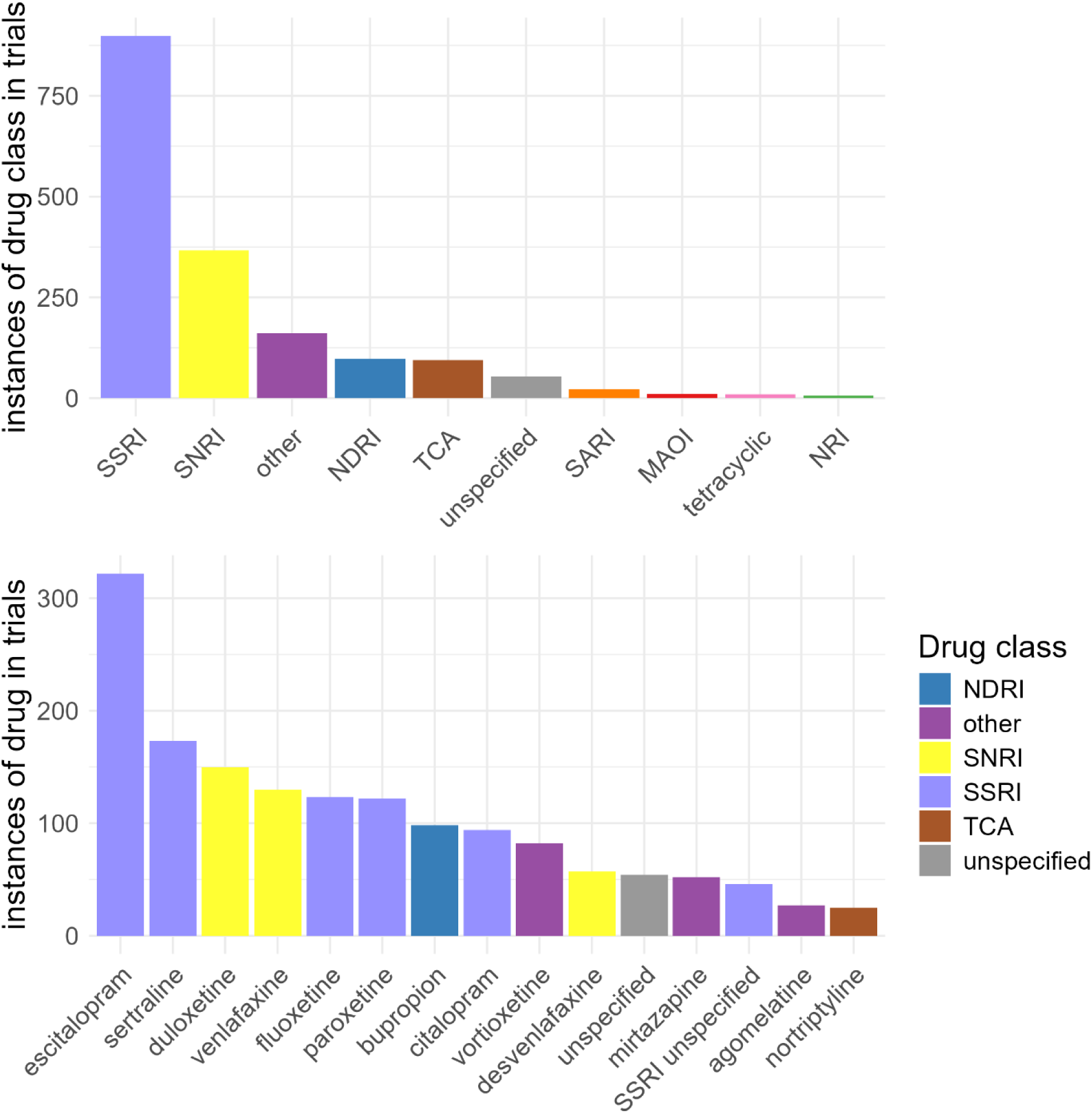
Distribution of drug interventions by antidepressant class (panel A) and antidepressant drug (panel B). In panel B, only drugs with >25 instances are included. MAOI: monoamine oxidase inhibitor; NDRI: norepinephrine–dopamine reuptake inhibitor; NRI: norepinephrine reuptake inhibitor; SARI: Serotonin antagonist and reuptake inhibitor; SNRI: serotonin-norepinephrine reuptake inhibitor; SSRI: selective serotonin reuptake inhibitor; TCA: tricyclic.

### Conditions

Within the 8,449 trials with categorised conditions, there were 14,394 instances of conditions, as some trials had several different conditions. Of these 14,394 conditions, most were classed in the mental and behavioural main category (n = 10,724, 74.9%). This was predominantly due to there being 7,631 (53.0%) instances of depression (Figure 5).

**Figure 5.**
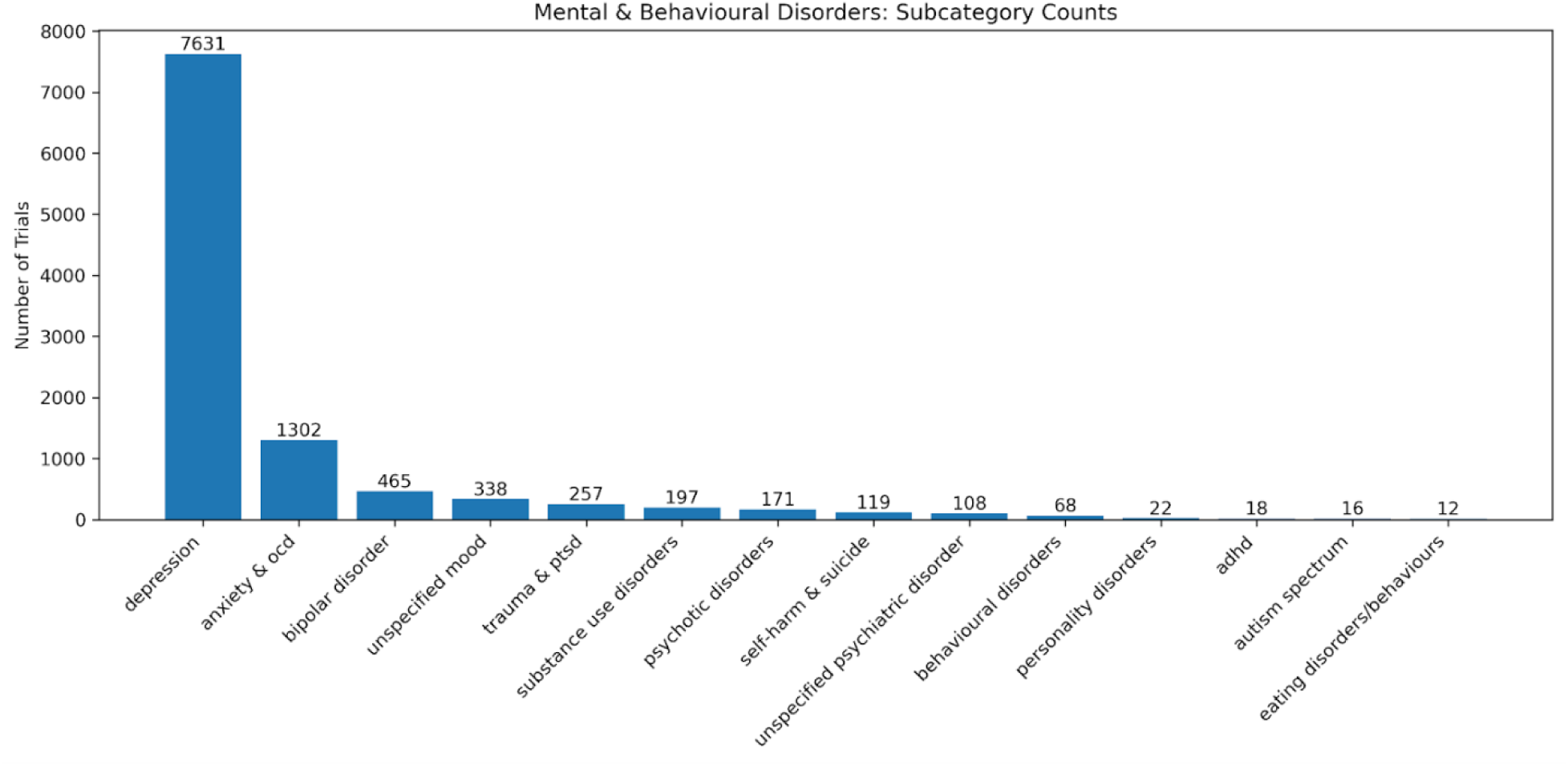
Number of trials containing conditions within the subcategories of the broader mental and behavioural main category.

Of 7,631 trials that investigated depression, 2,197 (28.8%) investigated other mental and behavioural disorders, with ‘anxiety & OCD’ being the most commonly co-examined subcategory (n = 1,264, 16.6%) (Figure 6). The ‘female & perinatal’ subcategory was the second most frequently examined in combination with depression (n = 530, 6.9%).

**Figure 6.**
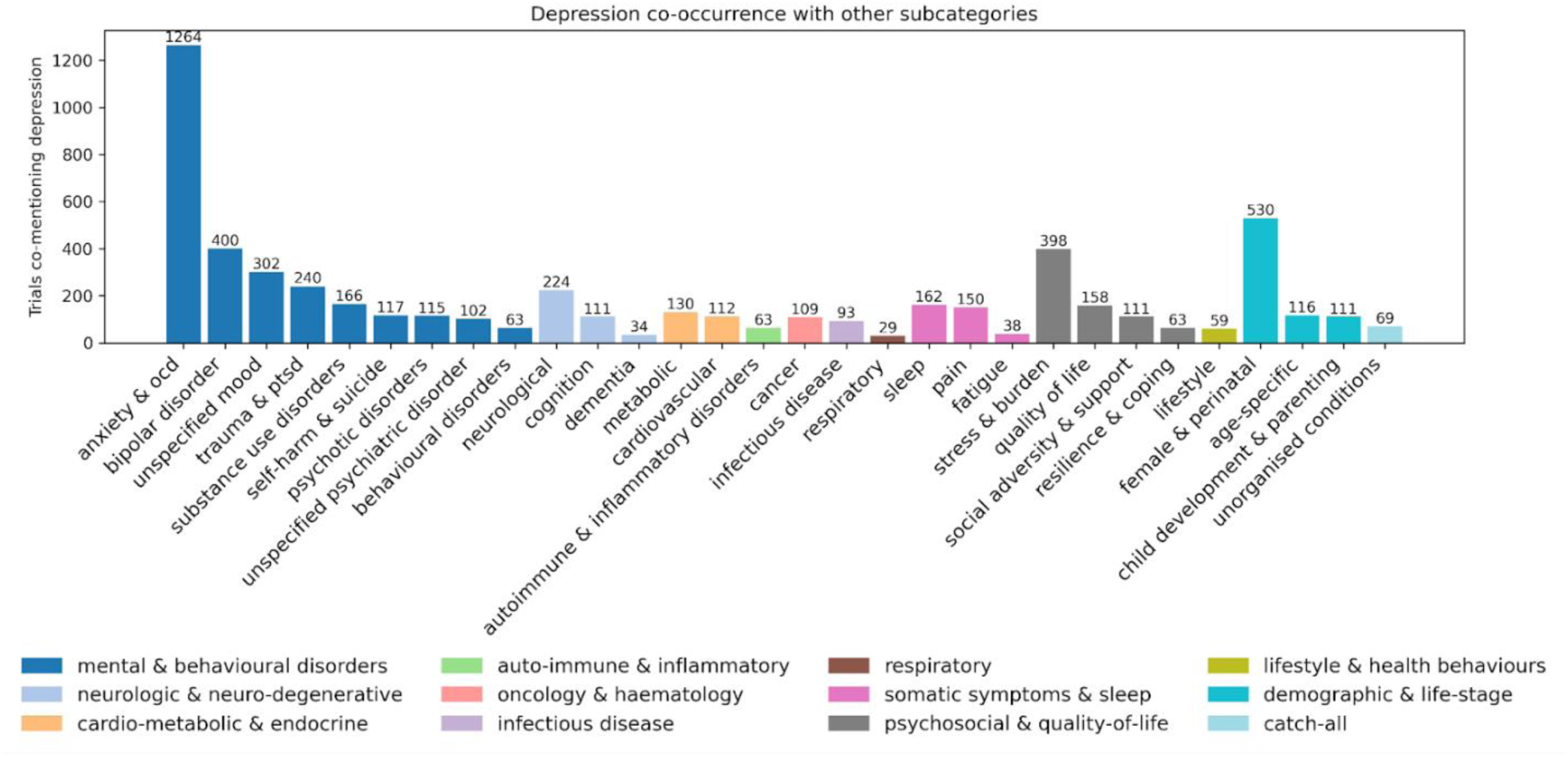
Co-occurrence of depression with other condition subcategories. Bars show the number of trials that mention depression and the listed subcategory at least once. Removed conditions with number of trials at 25 or below for readability.

### Temporal trends in pharmacological research

Trials into different classes of drugs to treat depression have changed substantially over time. Figure 7 shows the annual counts of trial arms containing SSRIs, SNRIs, NMDA receptor antagonists, and other classes from 1987 – 2024. Trials involving SSRIs rose sharply through the 1990s, peaked in 2009, and then declined. The number of trials studying NMDA receptor antagonist agents have increased since 2007.

**Figure 7.**
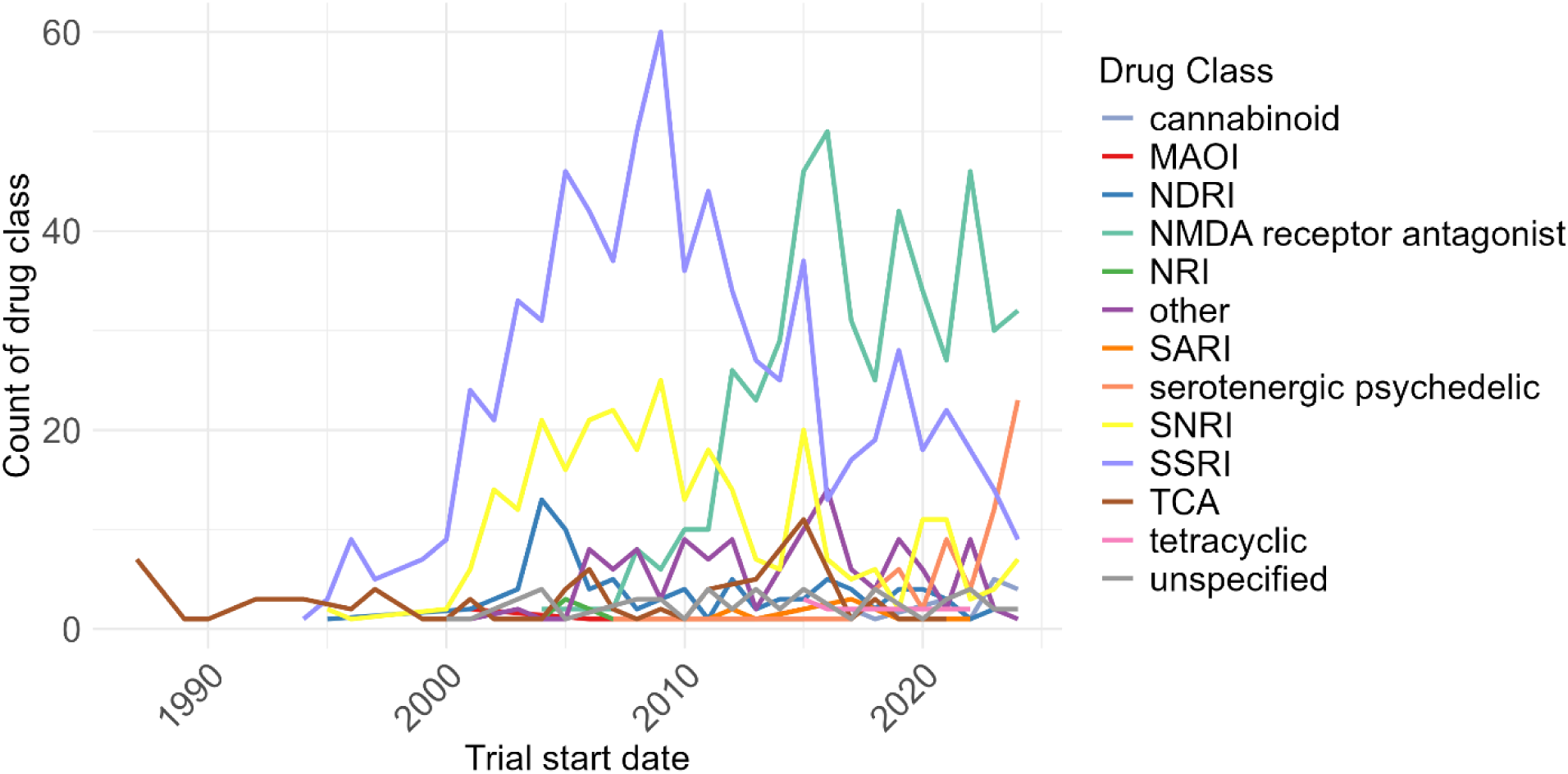
Trials over time, by drug class, 1987 to 2024. MAOI: monoamine oxidase inhibitor; NDRI: norepinephrine–dopamine reuptake inhibitor; NRI: norepinephrine reuptake inhibitor; SARI: Serotonin antagonist and reuptake inhibitor; SNRI: serotonin-norepinephrine reuptake inhibitor; SSRI: selective serotonin reuptake inhibitor; TCA: tricyclic.

### Biological sample availability

For each trial in ACT (n = 8853), vector scores were calculated based on query terms of specific text to aid with identifying trials that could be followed up for pharmacogenetic studies, for other biological omics studies, or to identify specific trial properties. Drug-related query groups tended to have higher mean scores and broader ranges than other query groups, suggesting that they could include a higher proportion of trials matching the search query. Distributions of five vector score terms (Biospecimens, Genetics, Proteomics, SSRIs and Antidepressant Response) are shown in Figure 8. These query groups were chosen to represent a mix of drug- and gene-related queries, as well as a range of score distributions and varying levels of specificity (e.g., broader terms like *biospecimens* versus narrower terms like *proteomics*).

**Figure 8.**
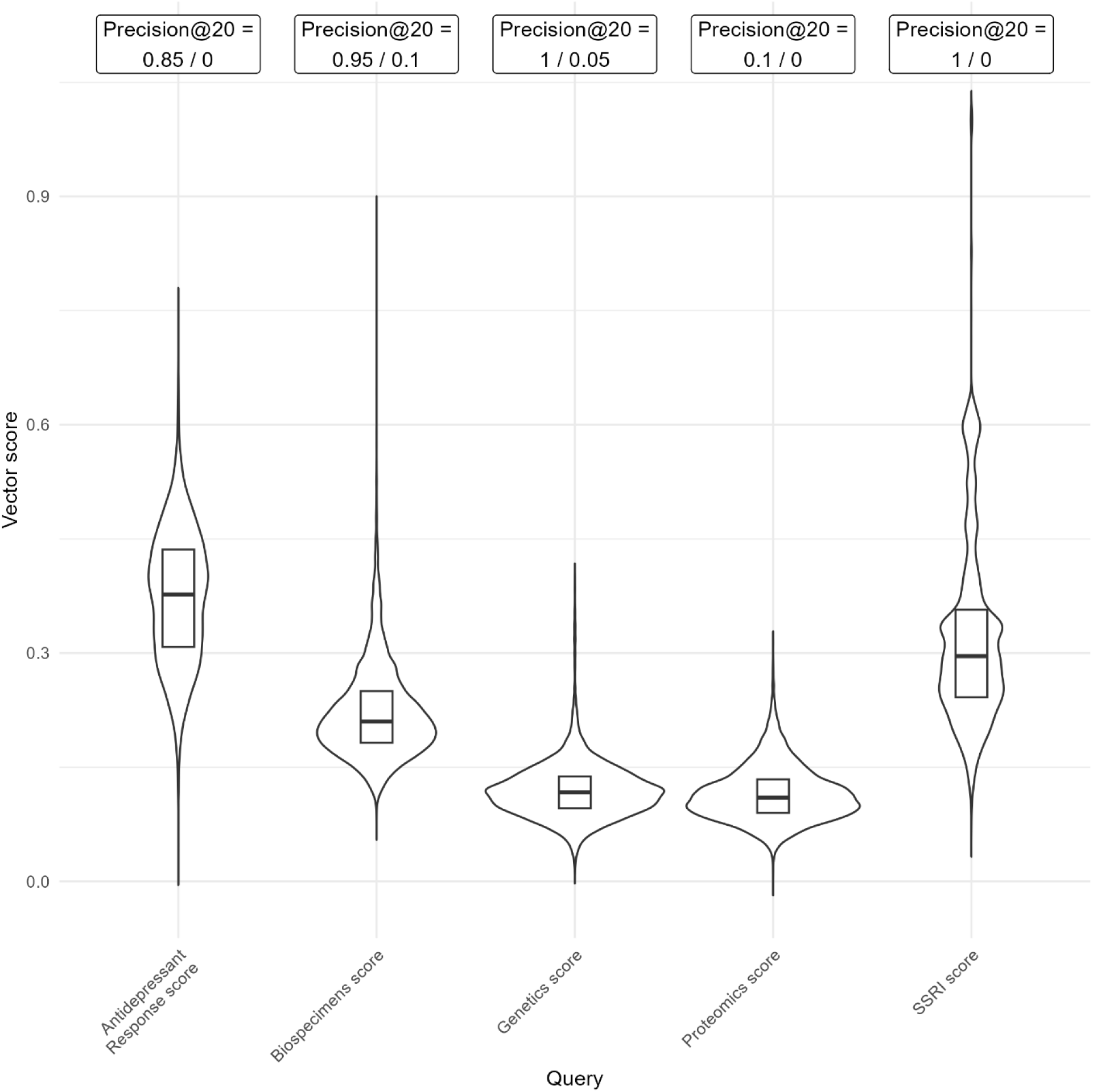
Distribution of vector scores calculated for five query terms of Biospecimens, Genetics, Proteomics, SSRIs (selective serotonin reuptake inhibitors) and Antidepressant Response, with precision values. Violin plots with embedded box plots show median (thick line), and inter-quartile range (thin line). Precision values show the proportion of trials with the relevant property, from the 20 trials with the highest and the lowest vector scores.

The precision at 20 highest and lowest values were calculated taken from classifying the 20 trials with the highest and lowest vector score in each term. Most query groups demonstrate high relevance among the top 20 results (95% - 100% precision) and low relevance among the lowest scores (0-10%), indicating their value as a search tool. However, where the overall range is narrow, as in the proteomics query, the top 20 results were only marginally more relevant than the bottom 20. This suggests that such scores are less effective for identifying relevant trials. Therefore, range and maximum value of scores must be considered when using in searches, with lower ranges and lower maximum scores being less useful in distinguishing relevant results. Distributions for all vector score terms are given in Supplementary figure 3.

### Strengths and limitations of the data

This data note describes a data set of clinical trials performed on depression, integrated across the US ClinicalTrials.gov (https://clinicaltrials.gov/) database, and the EU Clinical Trials Registry (https://www.clinicaltrialsregister.eu/), including trials from the inception of each resource until data download date of June 2024. The data set is a rich resource with information on trial design, interventions, sponsor, and information on publications reporting trial results. Trials are registered before data collection, and records may not be updated afterwards. Information on sample size, and the characteristics of trial participants is not given in the trial registers. Where possible we extracted information on sample sizes and participants’ demographic characteristics from publications, but this information is incomplete (see Supplementary Table 2, Supplementary Figure 1), and publications are not linked to the EU Clinical Trials Registry. A comprehensive analysis of trial participation by gender or race/ethnicity cannot, therefore, be performed. The data summary provided here focuses on trials assessing drugs used to treat depression, but the resource also provides a rich resource on trials using behavioural and digital interventions. The lack of sample size information for many trials is a key limitation. Of the 2218 studies with sample size information, 1281 (57.8%) had sample sizes of ≥100 participants and 242 (10.9%) of ≥1000 participants.

## Data Availability

This data set is openly available at https://kcl.figshare.com/articles/dataset/_b_Clinical_trials_in_depression_Integrated_collection_across_EU_and_US_registries_b_/30217441/0. Data fields are as described in Table 2. Data are available under the terms of the Creative Commons Attribution 4.0 International license (CC-BY 4.0).

Python and R scripts used for data analysis are also available at: https://github.com/louisesophieschindler/Clinical-trials-in-depression and https://github.com/katemstewart/Clinical_Trials_in_Depression

## Extended data

Supplementary information and material are available at https://kcl.figshare.com/articles/dataset/_b_Clinical_trials_in_depression_Integrated_collection_across_EU_and_US_registries_b_/30217441/0. Data are available under the terms of the Creative Commons Attribution 4.0 International license (CC-BY 4.0).

## Ethics

This study was an analysis of open access data from websites of two clinical trial repositories, and ethical permission is not required.

## Patient and Public Involvement

Patients and the public were not involved in the data extraction or summary.

## Grant information

This research was funded by Wellcome Mental Health Award (226770/Z/22/Z). The research was also part-funded by the National Institute for Health and Care Research (NIHR) Maudsley Biomedical Research Centre (BRC). The views expressed are those of the author(s) and not necessarily those of the NIHR or the Department of Health and Social Care. AMM is also supported by UKRI Grant MR/Z000548/1.

## Competing Interests

RB and SR were employees of Medicines Discovery Catapult when the research was performed. CML sits on the Scientific Advisory Board of Myriad Neuroscience.

## Acknowledgements

During the preparation of this work, the authors used ChatGPT (GPT-3.5/GPT-4) and Microsoft Copilot (GPT-4) to develop and troubleshoot sections of the code used during analysis, annotate scripts for reproducibility and clarity, and to rephrase text for clarity in the data note text. ChatGPT was used in the initial search of methods to evaluate information retrieval. After using these tools, the authors reviewed and edited the content as needed and take full responsibility for the content of the publication.

