## Supplementary Materials for "Clinical trials in depression: Integrated collection across EU and US registries"

Stewart K, Schindler L, et al.

### Supplementary Methods

Depression trials were identified by matching the conditions table to a regular expression based on the Depression Trial MeSH terms listed below. The set of conditions associated with the trials returned by the query were manually validated and irrelevant conditions were discarded. This refined condition set was used as a filter to identify depression trials, including those investigating conditions alongside depression (e.g. cancer and depression).

#### Depression Trial MeSH terms

1. depression
2. depressive disorder
3. depressive disorder, major
4. depression, postpartum
5. depressive disorder, treatment-resistant

#### Depression Trial terms

| | |
|---|---|
| 1 | depression |
| 2 | major depressive disorder |
| 3 | depressive symptoms |
| 4 | depressive disorder |
| 5 | depressive disorder, major |
| 6 | major depression |
| 7 | depression, anxiety |
| 8 | postpartum depression |
| 9 | treatment resistant depression |
| 10 | bipolar depression |
| 11 | depression, postpartum |
| 12 | unipolar depression |
| 13 | perinatal depression |
| 14 | major depressive disorder (mdd) |
| 15 | major depressive episode |
| 16 | anxiety depression |
| 17 | depressive disorder, treatment-resistant |
| 18 | depression, unipolar |
| 19 | depression, bipolar |
| 20 | post partum depression |
| 21 | postnatal depression |
| 22 | depressive episode |
| 23 | treatment-resistant depression |
| 24 | depression in old age |
| 25 | maternal depression |
| 26 | depressive disorders |
| 27 | post-stroke depression |
| 28 | depression in adolescence |

| | |
|---|---|
| 29 | depression mild |
| 30 | persistent depressive disorder |
| 31 | severe depression |
| 32 | moderate depression |
| 33 | depression moderate |
| 34 | adolescent depression |
| 35 | late life depression |
| 36 | major depressive disorders |
| 37 | post-partum depression |
| 38 | depressive disorder and anxiety disorders |
| 39 | antenatal depression |
| 40 | major depression with psychotic features |
| 41 | clinical depression |
| 42 | depression severe |
| 43 | psychotic depression |
| 44 | major depressive disorder, recurrent |
| 45 | treatment resistant major depressive disorder |
| 46 | geriatric depression |
| 47 | chronic depression |
| 48 | anxiety and depression |
| 49 | childhood depression |
| 50 | depression/anxiety |
| 51 | mental depression |
| 52 | atypical depression |
| 53 | bipolar i depression |
| 54 | mild depression |
| 55 | recurrent depressive disorder |
| 56 | severe major depression with psychotic features |
| 57 | recurrent depression |
| 58 | depression and suicide |
| 59 | suicide and depression |
| 60 | menopausal depression |
| 61 | treatment resistant depressive disorder |
| 62 | depression anxiety disorder |
| 63 | major depressive disorder, single episode, unspecified |
| 64 | depression symptoms |
| 65 | adjunctive treatment of major depressive disorder |
| 66 | prenatal depression |
| 67 | late-life depression |
| 68 | depressive state |
| 69 | postoperative depression |
| 70 | major depressive disorder, recurrent, in remission |
| 71 | bipolar disorder depression |
| 72 | perimenopausal depression |

| | |
|---|---|
| 73 | major depression disorder |
| 74 | major depressive disorder, recurrent, unspecified |
| 75 | depression, teen |
| 76 | unipolar major depression |
| 77 | major depressive disorder with psychotic features |
| 78 | therapy-resistant depression |
| 79 | resistant depression, treatment |
| 80 | mild to moderate depression |
| 81 | chronic depressive disorder |
| 82 | anxious depression |
| 83 | depressive disorder nos |
| 84 | subthreshold depression |
| 85 | antepartum depression |
| 86 | major depressive disorder patients |
| 87 | acute depression |
| 88 | chronic major depressive disorder |
| 89 | manic depression |
| 90 | depression not otherwise specified |
| 91 | depressive episodes, bipolar i depression |
| 92 | recurrent depressive disorder, current episode moderate |
| 93 | anxiety depression disorder |
| 94 | severe major depression |
| 95 | severe postpartum depression |
| 96 | resistant major depression |
| 97 | depression nos |
| 98 | depressive disorder not otherwise specified |
| 99 | minor depression |
| 100 | depressive disorder in mothers |
| 101 | depressive symptomatology |
| 102 | subclinical depressive symptoms |
| 103 | mild to moderate depressive symptoms |
| 104 | depression chronic |
| 105 | treatment resistant depression (trd) |
| 106 | minor depressive disorder |
| 107 | current major depressive disorder |
| 108 | symptoms of depression |
| 109 | anxiety depression (mild or not persistent) |
| 110 | outpatients / inpatients with depression |
| 111 | peripartum depression |
| 112 | winter depression |
| 113 | depressive disorder/psychology |
| 114 | depressive syndrome |
| 115 | recurrent major depression |
| 116 | depression in parkinson''s disease |

| | |
|---|---|
| 117 | major depressive disorder in pregnancy |

**Supplementary Table 1:** Vector search query groups and queries

| Query group | Queries |
|---|---|
| Blood Sample | blood sample<br>blood test |
| Biospecimens | biospecimens<br>cells<br>body fluid<br>sweat<br>saliva<br>urine<br>blood<br>tissue<br>solid tissue |
| Genetics | genetics<br>genotype<br>genotyping |
| Genome | genome<br>genomewide<br>genome-wide |
| Pharmacogenetics | pharmacogenetics |
| Epigenomics | epigenomics |
| Cytochrome P450 | CYP2D6<br>CYP2C19<br>cytochrome<br>P450 |
| Single Nucleotide Polymorphism | single nucleotide polymorphism<br>SNP<br>nucleotide |
| DNA | DNA |
| DNA methylation | DNA methylation |
| Proteomics | proteomics<br>protein expression |
| SSRI | SSRI<br>selective serotonin reuptake inhibitors |
| SNRI | SNRI<br>serotonin norepinephrine reuptake inhibitor |
| TCA | TCA<br>tricyclic antidepressants |
| NARI | NARI<br>norepinephrine reuptake inhibitor |
| MAOI | MAOI<br>monoamine oxidase inhibitor |
| Adverse Events/Side Effects | adverse events<br>side effects |
| Antidepressant Response | antidepressant response |
| Antidepressant Mechanisms | antidepressant mechanisms |
| Causal Inference | causal inference |

| | |
|---|---|
| Electronic Health Records | electronic health records<br>EHR<br>electronic medical records<br>EMR |
| Lived Experience | lived experience<br>PTSD<br>grief |
| Patient Involvement | patient involvement |

**Supplementary table 2:** Number of trials from which sample size and demographic data were extracted from publications following trial completion. Where information was extracted from multiple PMIDs, the mode is reported.

| variable | no. trials with available data |
|---|---|
| trial id | 8,853 |
| sample size mode | 1,670 |
| sample size mean | 2,218 |
| age mean mode | 793 |
| age mean mean | 941 |
| age minimum mode | 5,212 |
| age minimum mean | 5,394 |
| age maximum mode | 3,288 |
| age maximum mean | 3,450 |
| bmi mean mode | 143 |
| bmi mean mean | 158 |
| bmi minimum mode | 329 |
| bmi minimum mean | 335 |
| bmi maximum mode | 374 |
| bmi maximum mean | 381 |
| male count mode | 785 |
| male count mean | 944 |
| female count mode | 951 |
| female count mean | 1,145 |
| ethnicity counts | 713 |

| | |
|---|---|
| American Indian or Alaska Native count mode | 78 |
| American Indian or Alaska Native count mean | 79 |
| Asian count mode | 210 |
| Asian count mean | 226 |
| Black or African American count mode | 341 |
| Black or African American count mean | 393 |
| Hispanic or Latino count mode | 270 |
| Hispanic or Latino count mean | 319 |
| Native Hawaiian or Pacific Islander count mode | 30 |
| Native Hawaiian or Pacific Islander count mean | 30 |
| White count mode | 444 |
| White count mean | 532 |
| More than one race count mode | 82 |
| More than one race count mean | 87 |
| Other count mode | 174 |
| Other count mean | 199 |

**Supplementary Table 3:** Distribution of category of interventions included in trials. Note that many trials had multiple interventions within trial arms or across trial arm (for example a drug intervention with and without psychological treatment), and all interventions are included.

| Name | Count |
|---|---|
| Behavioural | 3,291 |
| Biological | 48 |
| Combination | 33 |
| Device | 986 |
| Diagnostic | 33 |
| Dietary | 157 |
| Drug | 3,659 |
| Genetic | 14 |
| Other | 1,395 |
| Procedure | 205 |
| Radiation | 11 |

**Supplementary Table 4:** Number of trials sponsored by specific institutions or companies, for top 10 sponsors in the categories of all trials, trials with a drug intervention, and trials with a drug to treat depression.

| All trials | | Trials including a drug intervention | | Trials including a drug to treat depression | |
|---|---|---|---|---|---|
| Sponsor | count | Sponsor | count | Sponsor | count |
| Massachusetts General Hospital | 172 | Eli Lilly and Company | 80 | Eli Lilly and Company | 54 |
| University of Pittsburgh | 98 | Massachusetts General Hospital | 74 | New York State Psychiatric Institute | 44 |
| New York State Psychiatric Institute | 93 | GSK | 62 | H. Lundbeck A/S | 35 |
| VA Office of Research and Development | 87 | New York State Psychiatric Institute | 56 | Massachusetts General Hospital | 35 |
| Stanford University | 85 | National Institute of Mental Health (NIMH) | 48 | GSK | 29 |
| University of California, Los Angeles | 82 | H. Lundbeck A/S | 47 | University of Pittsburgh | 24 |
| Eli Lilly and Company | 81 | Janssen Research & Development, LLC | 40 | Wyeth is now a wholly owned subsidiary of Pfizer | 23 |
| Centre for Addiction and Mental Health | 70 | AstraZeneca | 34 | Janssen Research & Development, LLC | 21 |
| Washington University School of Medicine | 66 | University of Oxford | 30 | Yale University | 20 |
| Weill Medical College of Cornell University | 65 | University of Pittsburgh | 30 | Washington University School of Medicine | 17 |

**Supplementary Figure 1:** Missingness of demographic data from 8,853 trials, showing number of trials with missing data.

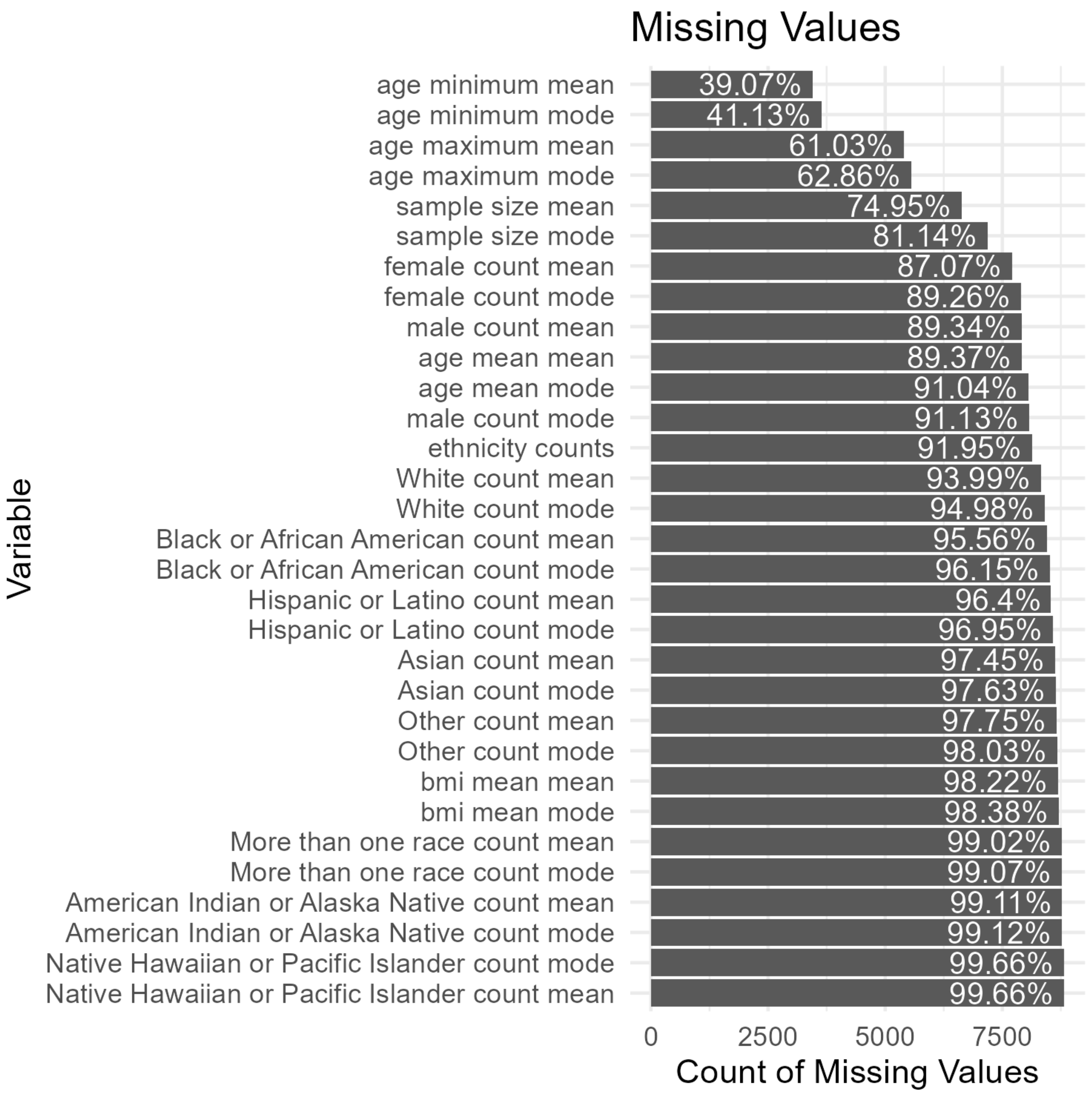

**Supplementary Figure 2:** Distribution of Emerging Depression Interventions (EDIT) including NMDA receptor antagonists, psychedelics, and cannabinoids by class (panel A) and by drug (panel B).

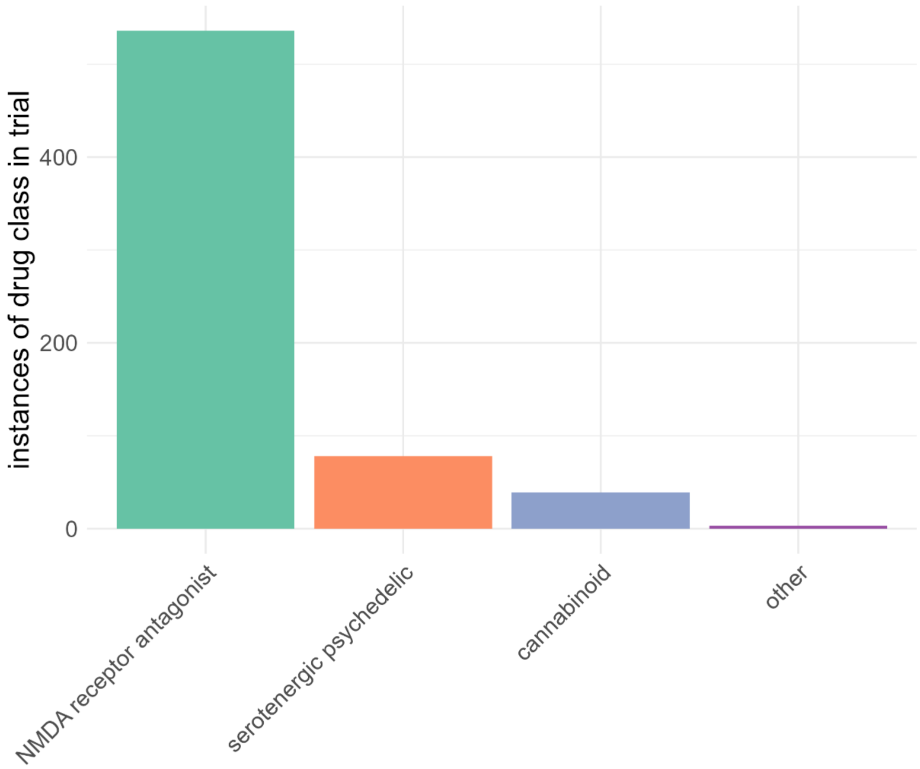


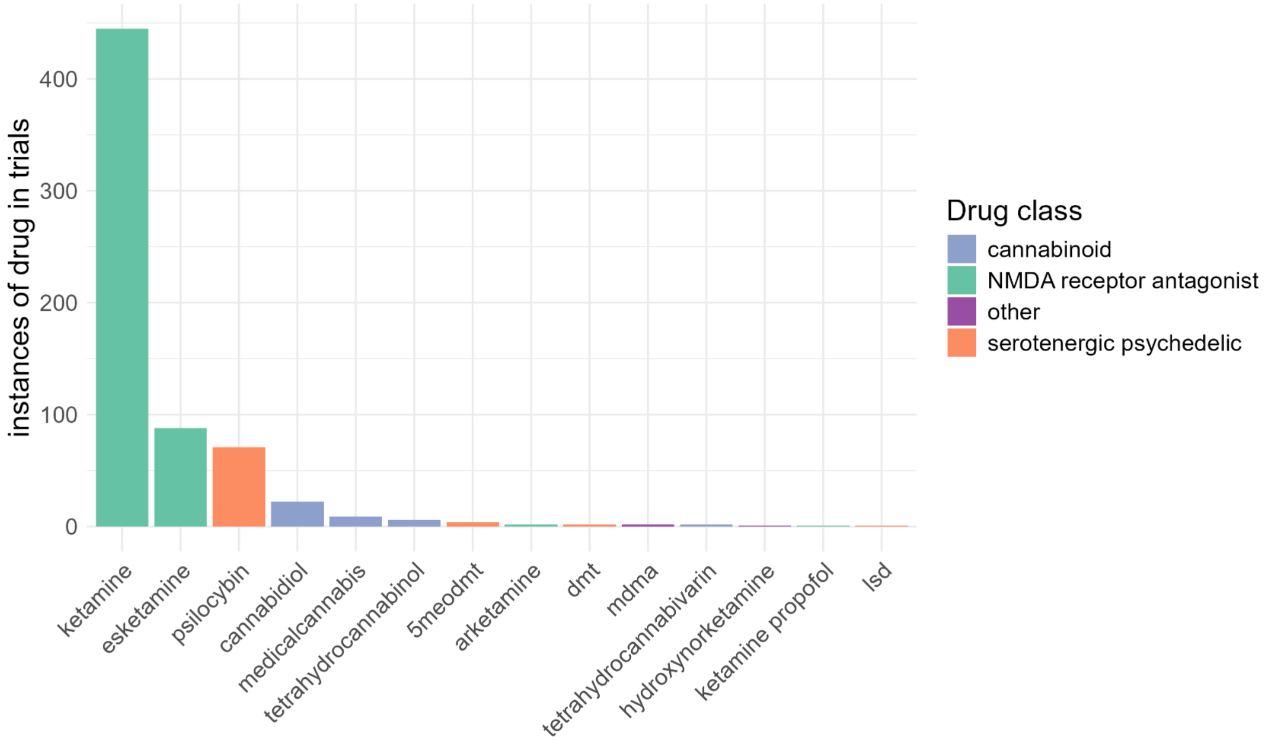

**Supplementary Figure 3:** Distribution of vector scores for trial characteristics. Violin plots show median (thick line), and inter-quartile range (thin line). All clinical trials (n = 8853) were assigned a vector score for each query. MAOI: monoamine oxidase inhibitor; NARI: norepinephrine reuptake inhibitor; SNRI: serotonin-norepinephrine reuptake inhibitor; SSRI: selective serotonin reuptake inhibitor; TCA: tricyclic.

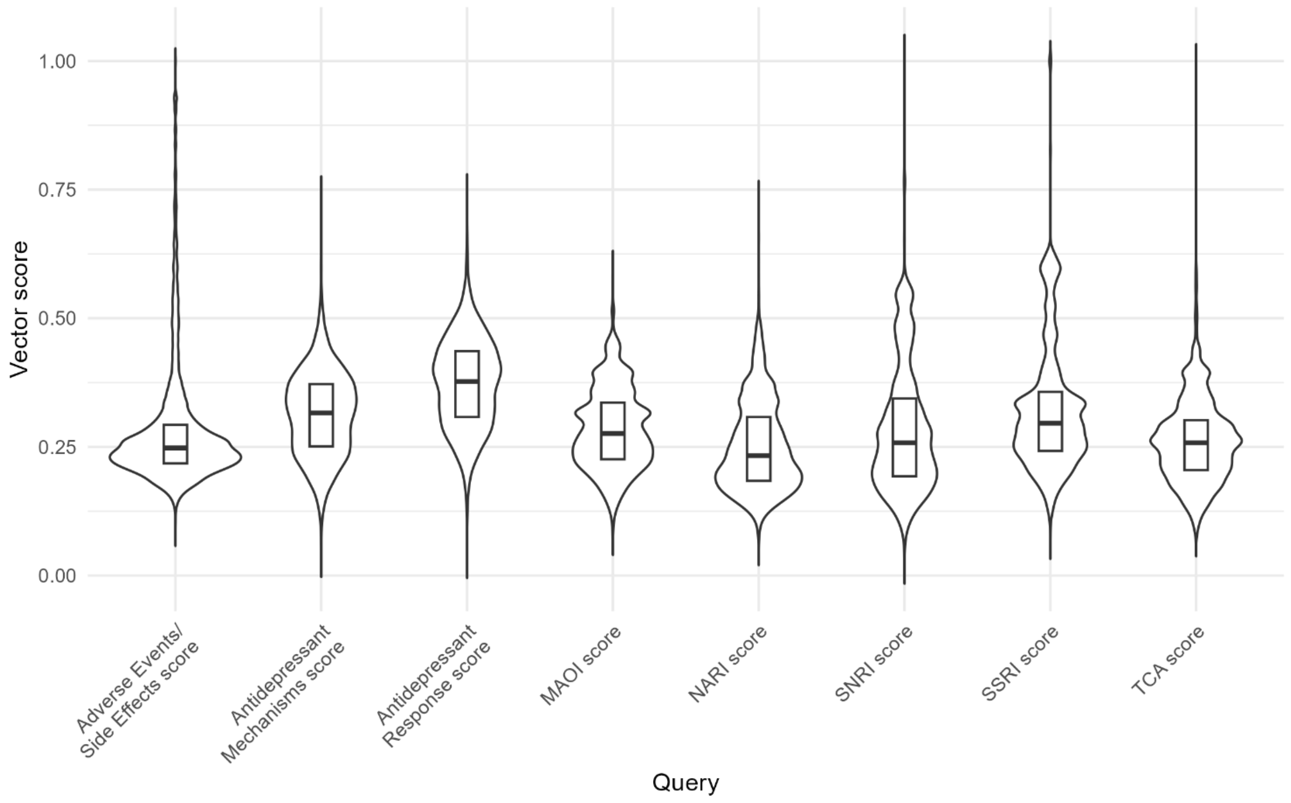

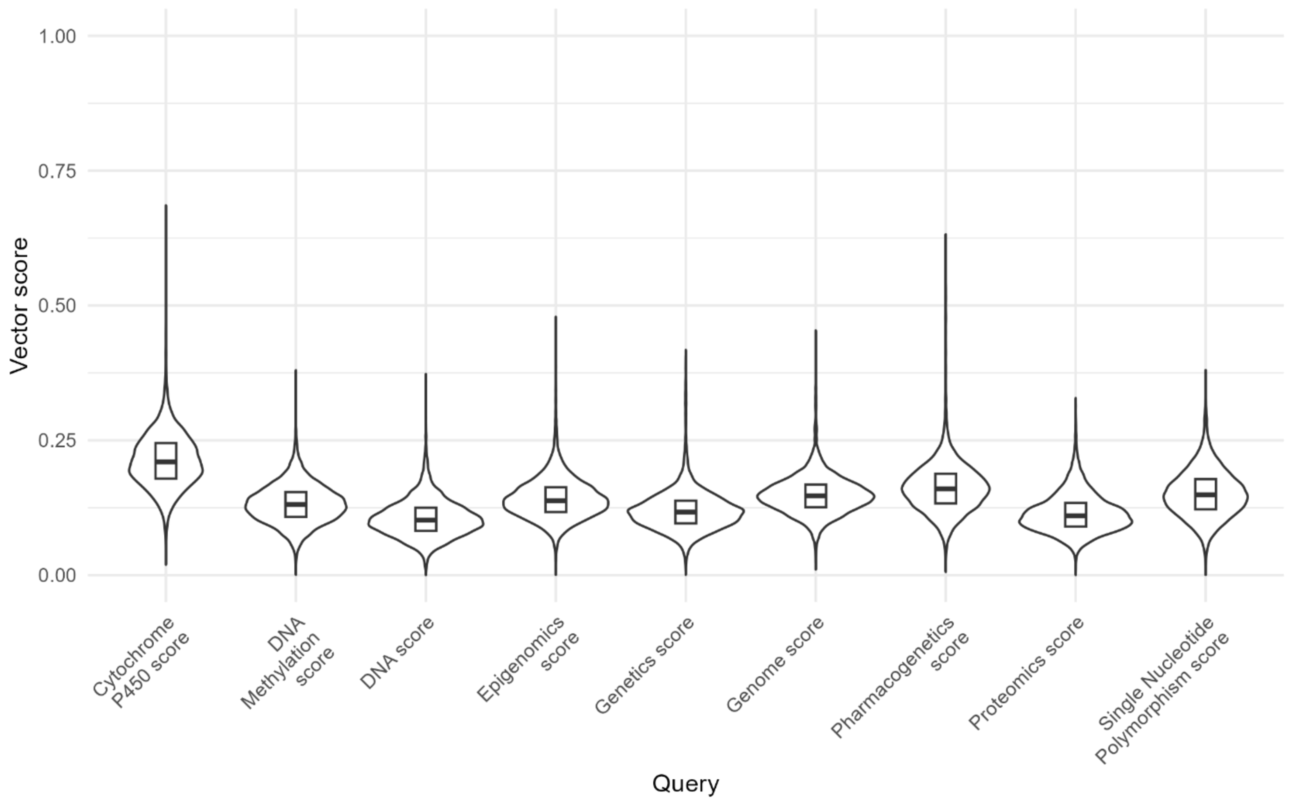
1.00
0.75
0.50
0.25
0.00
Vector score
Cytochrome P450 score
DNA Methylation score
DNA score
Epigenomics score
Genetics score
Genome score
Pharmacogenetics score
Proteomics score
Single Nucleotide Polymorphism score
Query

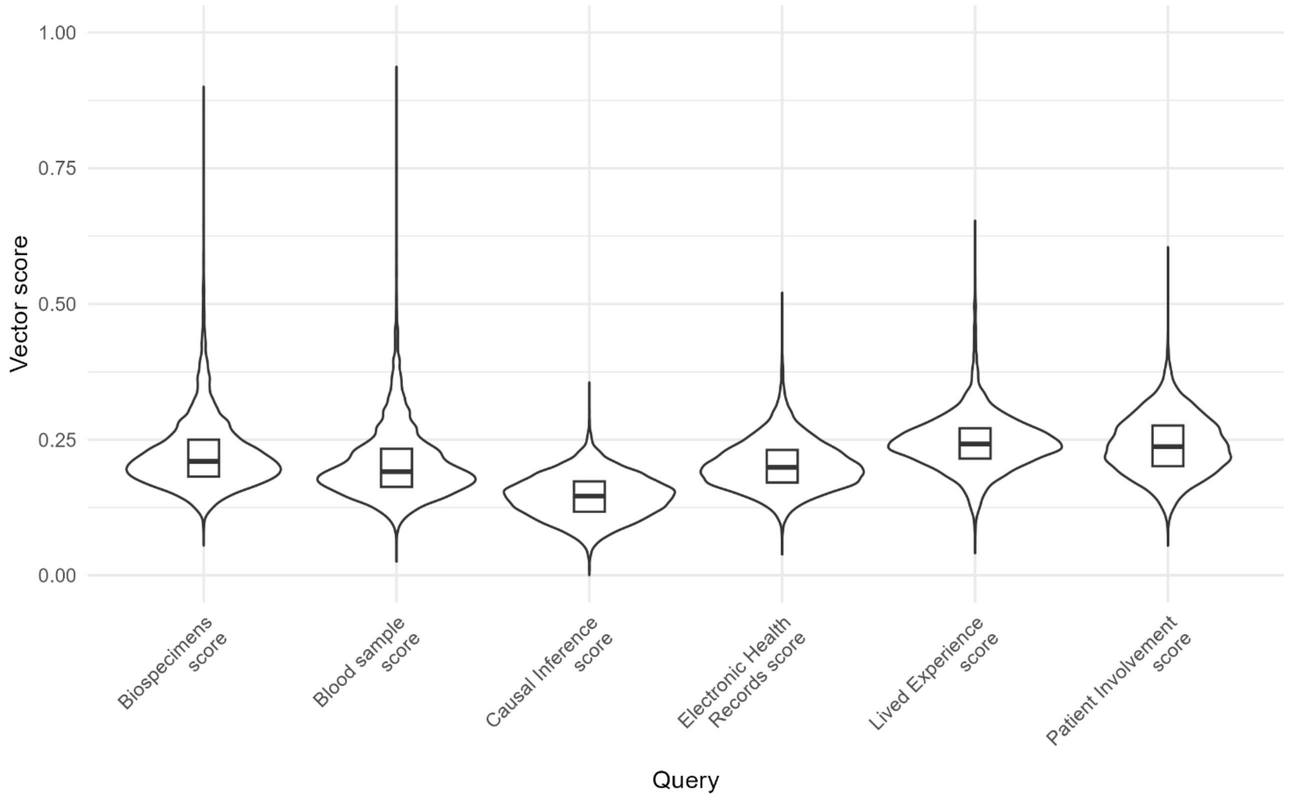
1.00
0.75
0.50
0.25
0.00
Vector score
Biospecimens score
Blood sample score
Causal Inference score
Electronic Health Records score
Lived Experience score
Patient Involvement score
Query
